# Exploring the feasibility, acceptability and impact of genomic newborn screening for rare diseases in England: A study protocol for the Generation Study - Process and Impact Evaluation (Version 2)

**DOI:** 10.1101/2024.05.14.24307295

**Authors:** Celine Lewis, Emma Beecham, Felicity Boardman, Gráinne Brady, James Buchanan, Sigrún Clark, Katie Gilchrist, Pia Hardelid, Amy Hunter, Jennifer Jones, Kerry Leeson-Beevers, Bethany Stafford-Smith, Joachim Tan, Cecilia Vindrola-Padros, Martin Vu, Wing Han Wu, Ania Zylbersztejn, Melissa Hill

**Affiliations:** Population, Policy and Practice, UCL Great Ormond Street Institute of Child Health, London, UK; Rapid Research Evaluation and Appraisal Lab (RREAL), University College London, London, UK; Warwick Medical School, University of Warwick, Coventry, UK; Health Economics and Policy Research Unit (HEPRU), Queen Mary University of London, London, UK; National Institute for Health and Care Research Barts Biomedical Research Centre, London, UK; Genetic Alliance UK, London, UK; Department of Population Health Sciences, University of Leicester, Leicester, UK; Alström Syndrome UK and Breaking Down Barriers, Torquay, UK; NHS North Thames Genomic Laboratory Hub, Great Ormond Street Hospital for Children NHS Foundation Trust, London, UK; Genetics and Genomic Medicine, UCL Great Ormond Street Institute of Child Health, London, UK

## Abstract

The role of genomics in healthcare is expanding rapidly and many countries are exploring the possibility of using genomic sequencing to expand current newborn screening programmes. Offering routine genomic newborn screening (gNBS) would allow newborn screening to include a much broader range of rare conditions, but there are many technical, practical, psychosocial, ethical and economic challenges to be addressed. Genomics England and NHS England have established the Generation Study to deliver gNBS for 100,000 births to explore the benefits, challenges, and practicalities of offering gNBS to parents in England. Here we describe the study protocol for the Generation Study - Process and Impact Evaluation, an independent mixed-methods evaluation of the Generation Study. The evaluation will have oversight from a Study Advisory Group that includes academic, clinical and patient representatives and a Patient and Public Involvement and Engagement (PPIE) Advisory Group that includes members from parent and patient organisations and parents with relevant experiences. The Process and Impact Evaluation will examine whether offering gNBS in routine care is feasible and acceptable and inform our understanding of the clinical utility and cost effectiveness of gNBS in England. Through surveys and interviews conducted at two time points we will explore the attitudes and experiences of parents, professionals and patient organisations. We will also consider the clinical, psychosocial and health economic impacts, both positive and negative. The results will be presented at national and international conferences and submitted for peer review and publication.

## SUMMARY

The Generation Study - Process and Impact Evaluation aims to provide an independent mixed-methods evaluation of the use of genomic newborn sequencing (gNBS) for the early diagnosis of rare, childhood-onset, actionable genetic conditions.

The evaluation comprises seven core studies:

### Study 1: Identifying goals, challenges and early lessons in implementation

Interviews at outset of the Generation Study with professionals who are the Generation Study’s “designers” and “early implementors”; shadowing of processes and pathways; and documentary analysis. Interviews towards the end of Generation Study recruitment with professionals including the “designers” and “early implementers” and others who have become involved over time.

### Study 2: Impact, experiences and attitudes of parents

Survey and follow-up interviews with parents following receipt of gNBS results (Time 1) and 12 months later (Time 2) to assess acceptability, experience, attitudes and impact (positive and negative).

### Study 3: Gathering wider professional viewpoints about the Generation Study and gNBS implementation

Survey with staff delivering the Generation Study at early adopter sites and professionals from a range of relevant backgrounds across England.

### Study 4: Views of the rare disease community

Surveys and interviews with advocates from rare disease support groups to examine attitudes to gNBS including positive and negative impacts as well as unintended consequences on parents and the rare disease community more broadly.

### Study 5: Public views

Survey conducted with a market research company to capture views of with a diverse group of parents in terms of age, gender, ethnicity, socio-economic status and geographical location to evaluate the key findings around acceptability and attitudes towards gNBS with parents who have not been involved in the Generation Study.

### Study 6: Cost effectiveness evaluation

Assessment of the impact of gNBS on healthcare resource use and costs, quality of life outcomes and non-health-related outcomes, to inform the Genomics England health economics model.

### Study 7: Clinical utility assessment

Assessment of the clinical utility of gNBS for health-related outcomes by comparing hospital contact and mortality rates in various groups of children (confirmed diagnosis, population controls), making use of the longitudinal healthcare data linked to the Generation Study and control groups primarily drawn from ECHILD.

Summary Table: Research Questions, Methods and Outputs.

**Summary Table: Research Questions, Methods and Outputs**
| Work | Research questions | Data collection | Data analysis | Expected output |
| --- | --- | --- | --- | --- |
| Scoping |  | Meetings with key stakeholders<br>Review of documents | Rapid qualitative data analysis. | Determine scope and agree priority research questions and methods. Stakeholder engagement. |
| Study 1: Understanding goals and challenges | Is the use of WGS as a tool for early diagnosis of rare, childhood-onset, actionable genetic conditions feasible and acceptable to professionals?<br>What are the implementation processes being used to deliver the Generation Study and how do these vary across sites and over time?<br>What are the effective approaches to implementation for delivery of gNBS including identified barriers and facilitators?<br>What are professionals' experiences of and attitudes towards delivering gNBS?<br>What are professionals' education and training needs to support delivery of gNBS?<br>What factors impact access and uptake to the Generation Study? | In depth interviews with designers and early implementers at two time points<br>Observations of local recruitment processes<br>Documentary analysis<br>Exit survey | Rapid qualitative data analysis<br>In-depth framework analysis | Slide presentations, infographic and report on interim findings<br>Final report |
| Study 2: Mixed methods study with parents | Is the use of WGS as a tool for early diagnosis of rare, childhood-onset, actionable genetic conditions feasible and acceptable to parents?<br>What are parents' experience of joining the Generation Study?<br>Did parents perceive that information about gNBS was effectively conveyed and that they were able to make an informed choice? | Survey with parents after receipt of gNBS result<br>In-depth interviews with a subset of survey participants | Descriptive and inferential statistics<br>Rapid qualitative data analysis. | Slide presentations, infographic and report on interim findings<br>Final report |
|  | What factors impact access and uptake to gNBS?<br>What are parents' information and support needs to facilitate decision making, receiving results and ongoing care and support networks?<br>Can findings be returned effectively using the approach adopted for the study overall?<br>What are parents' experiences of receiving a 'no condition suspected' gNBS result?<br>What are parents' experiences of receiving a positive gNBS result, including the downstream pathways for those with a true or false positive result?<br>What are the wider positive and negative impacts on families including parental mental health?<br>What are parents' experiences of receiving either a 'no condition suspected' or a positive gNBS result over the longer-term? |  |  |  |
| Study 3: Wider viewpoints of professionals | Is the use of gNBS as a tool for early diagnosis of rare, childhood-onset, actionable genetic conditions feasible and acceptable to the wider workforce?<br>What are wider workforces' experiences and attitudes to the use of gNBS as a tool for early diagnosis of rare, childhood-onset, actionable genetic conditions?<br>What are the downstream implications of offering gNBS including impacts on the wider clinical workforce? | Survey with professionals | Descriptive statistics / thematic analysis | Slide presentations<br>Final report |
| Study 4: Views of rare disease community | How prepared are patient organisations to support families taking part in gNBS? | Survey with rare disease support group representatives | Descriptive statistics / thematic analysis | Slide presentations on survey and interview findings |
|  | What impact do patient organisations think gNBS will have on the families they support now and in the future? | In-depth interviews with a subset of survey participants |  | Final report plus stand-alone report in Genetic Alliance UK branding |
| Study 5: Public views | What are parents from the general public (i.e. those who have not been invited to take part in the Generation Study) attitudes towards gNBS?<br>Do parents from different ethnic or educational backgrounds differ on their views of gNBS?<br>Do parents from the general public view gNBS as acceptable? | Survey with diverse parent groups through market research company | Descriptive and inferential statistics | Slide presentations on survey findings<br>Final report |
| Study 6: Cost effectiveness | What is the impact on secondary healthcare resource use including estimates of end-to-end costs – from recruitment, through sample processing, sequencing and interpretation through to clinical care?<br>What are the positive and negative impacts on health-related outcomes including: morbidity, mortality and quality of life?<br>What is the impact on non-health-related outcomes including: perceived personal utility and psychosocial wellbeing? | Survey measures with parents at consent and after receipt of gNBS results at two timepoints<br>Linked Hospital Episode Statistics data<br>Linked data on mortality and morbidity outcomes.<br>Resource utilisation for delivering gNBS as a service, with data collected during the initial period of the Generation Study and updated towards the end of the study.<br>Resource utilisation for the national scale-up of gNBS | Microcosting analysis<br>Descriptive and inferential statistics; Subgroup and threshold analyses<br>Quasi-experimental methods<br>Multiple imputation for missing data if required. | Slide presentations on interim findings<br>Final report |
| Study 7: Clinical utility assessment | What are the positive and negative impacts on health-related outcomes (morbidity, mortality)?<br>What is the prevalence of the conditions looked for in the newborn population? | Use of Genomics England cohort data linked to administrative data, ECHILD, additional data for controls. | Generalised linear models/survival analyses applied in the linked cohort data | Slide presentations on interim findings<br>Final report (month 24). |
|  | How many receive a 'condition suspected' results and how many would be expected if it were adopted nationally?<br>What is the level of uptake of the programme and what would the level of uptake of genomic newborn screening be if it were adopted nationally? |  |  | Code and methodology for future studies |
| Integration of findings |  |  | Integration of findings of the Process and Impact evaluations | Slide presentation and animations describing findings<br>Final report |

## AIM

To conduct a process and impact evaluation on the use of genomic newborn screening for the early diagnosis of rare conditions that can be treated in early childhood.

## PRIMARY RESEARCH QUESTIONS

1. Is the use of whole genome sequencing (WGS) as a tool for early diagnosis of rare, childhood-onset, actionable genetic conditions feasible and acceptable?
2. What is the impact (both positive and negative) of the programme on stakeholders and the wider system?
3. What are stakeholders’ experiences and attitudes to the use of WGS as a tool for early diagnosis of rare, childhood-onset, actionable genetic conditions?
4. What are the implementation processes being used to deliver the Generation Study and how do these vary across sites?

## RESEAERCH TEAM

Dr Celine Lewis, UCL Great Ormond Street Institute of Child Health

Professor Cecilia Vindrola-Padros, UCL Rapid Research Evaluation and Appraisal Lab

Ms Sigrún Clark, UCL Rapid Research Evaluation and Appraisal Lab

Ms Katie Gilchrist, UCL Rapid Research Evaluation and Appraisal Lab

Ms Ailey McCleod, UCL Rapid Research Evaluation and Appraisal Lab

Ms Grainne Brady, UCL Rapid Research Evaluation and Appraisal Lab

Dr Emma Beecham, UCL Rapid Research Evaluation and Appraisal Lab

Professor Felicity Boardman, University of Warwick

Dr James Buchanan, Queen Mary University of London

Dr Martin Vu, Queen Mary University of London

Professor Pia Hardelid, UCL Great Ormond Street Institute of Child Health

Dr Joachim Tan, UCL Great Ormond Street Institute of Child Health

Dr Ania Zylbersztejn, UCL Great Ormond Street Institute of Child Health

Dr Amy Hunter, Genetic Alliance UK

Dr Jennifer Jones, Genetic Alliance UK

Mrs Kerry Leeson-Beevers, Alström Syndrome UK and Breaking Down Barriers

Dr Melissa Hill, Great Ormond Street Hospital NHS Foundation Trust

Ms Bethany Stafford-Smith, Great Ormond Street Hospital NHS Foundation Trust

Ms Sweta Karthikeyan, Great Ormond Street Hospital NHS Foundation Trust

Mrs Wing Han Wu, Great Ormond Street Hospital NHS Foundation Trust

## STUDY OVERSIGHT

The Process and Impact Evaluation will have oversight from two advisory groups:

### Study Advisory Group

The Study Advisory Group, chaired by Dr Rachel Knowles (UCL Great Ormond Street Institute of Child Health), with members from academic, clinical and PPI backgrounds. PPI members will be from relevant patient organistations. The Study Advisory Group will oversee and guide the study. They will meet every quarter to review study progress and provide guidance.

### Patient & Public Involvement (PPI) Advisory Group

A PPI Advisory Group, chaired by Kerry Leeson-Beevers (Breaking Down Barriers), will include members representing relevant patient organisations and parent support groups and parent ambassadors with relevant experiences. The overarching goal of the PPI Advisory Group will be to ensure the preferences and priorities of parents are central to the research and are included through every stage. The PPI Advisory Group will meet every quarter to review study materials and approaches to recruitment and help interpret findings.

By having a separate PPI Advisory Group we hope to create a meeting space where parents can feel comfortable to share their views. During the study the PPI members of the Study Advisory Group and the PPI Advisory Group will be supported through one-to-one calls or emails to check-in with members after meetings. These interactions support members and also allow them to share or discuss items they didn’t feel comfortable bringing up in the group. Informal training will also be available, for example to explain newborn screening and the Generation Study.

## STUDY OVERVIEW

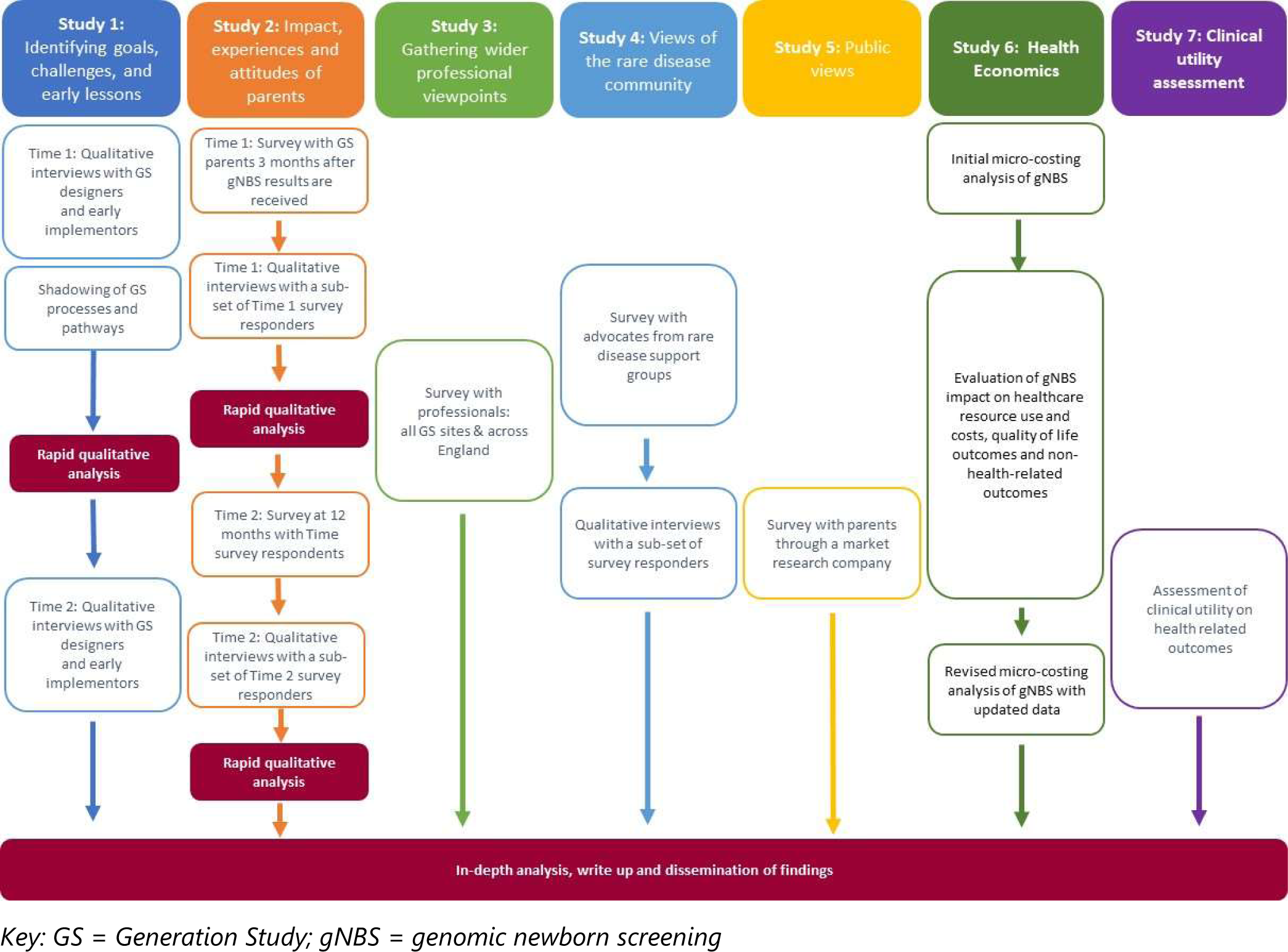

## RATIONALE

The Generation Study aims to explore the benefits, challenges, and practicalities of offering WGS to parents of newborns, to accelerate diagnosis and access to treatment for rare genetic conditions. The Process and Impact Evaluation has been funded by Genomics England to provide an independent mixed-methods evaluation of the use of gNBS for the early diagnosis of rare, childhood-onset, actionable genetic conditions. The evaluation will examine whether offering gNBS in routine care is feasible and acceptable and will inform our understanding of the clinical utility and cost effectiveness of offering gNBS. We will also explore the attitudes and experiences of parents who have been offered gNBS and consider the clinical and psychosocial impacts, both positive and negative.

## THEORETICAL FRAMEWORK

The evaluation will draw on several theoretical frameworks and measures. The use of theory-based and pre-specified constructs will help to generalise the findings and enable integration across the various studies, enabling us to build a stronger evidence base. For example, in order to understand the perspective of parents, we will use a number of parent reported outcome measures including decisional regret, and quality of life in our survey.

The philosophical approach that will underpin our mixed methods study design is pragmatism.^1, 2^ Pragmatism is a philosophical movement that states that an ideology or approach is true if it works satisfactorily. In that sense at its core it is problem-centred and puts the research question and generation of useful, actionable results above all else.^2^ It is frequently used in mixed methods research because it is focused on real-world issues and problems that need solving and has no loyalty to a specific philosophical stance. In that sense it permits different assumptions about worldviews and values both subjective and objective knowledge. Importantly, it allows the researchers to respond to the needs of the study as new evidence comes up, such as making methodological adjustments e.g. to research questions and/or data collection methods which will clearly be important given the fast turnaround of the project.

For the Process Evaluation we will use a theory driven approach to understand how the Generation Study is being implemented. We will draw on both the Consolidated Framework for Implementation Research (CFIR)^3^ and Normalisation Process Theory (NPT)^4–6^ when developing the interview topic guides and analysing the interview data. The CFIR can be used as an explanatory framework to systematically assess the contextual factors including barriers and facilitators that influence implementation and adoption,^3^ and has been used previously to evaluate implementation in other areas of genomic medicine.^7–10^ This framework is also well-suited to approaches involving rapid-cycle evaluation.^11^ The CFIR framework provides a taxonomy of operationally defined constructs that are likely to influence implementation of complex programs, organised into five major domains: 1) Intervention Characteristics (features of the intervention itself which might influence implementation e.g. complexity); 2) Outer Setting (features of the implementation organisation e.g. leadership engagement); 3) Inner Setting (features of the external context or environment e.g. readiness); 4) Characteristics of Individuals (e.g. knowledge and beliefs of individuals); and 5) Process (strategies or tactics which might influence implantation e.g. planning).

In addition, as this is the first time gNBS will be implemented within a public health setting on a large scale we will also draw on NPT, which is an implementation process theory that focuses on the work that individuals and groups do to enable an intervention to become routine or normalised.^4–6^ NPT emphasises agency, cooperation and coordination in a social system as key components of implementing complex interventions. NPT can be applied to explain how changes in the way people think about and use the intervention occur and identify factors that promote or inhibit the routine incorporation of the intervention into everyday practice. NPT proposes four constructs that represent different kinds of work that individuals and groups do around implementing a new practice: Coherence, Cognitive Participation, Collective Action, and Reflexive Monitoring. NPT will be used to guide the development of survey and interview questions and to inform the analysis of qualitative interview data.

## STUDY DESIGN

This study will use a longitudinal mixed methods research design using qualitative and quantitative approaches for data collection. Mixed methods approaches are frequently used to characterise complex healthcare systems as comparing the results of multiple data sets provides a more complete understanding of the topic.^12^ To deliver this evaluation with formative feedback, we will draw on the rapid feedback and rapid cycle evaluations used in the Rapid Research Evaluation And Appraisal Lab (RREAL).^13, 14^

Our evaluation comprises an initial scoping study and seven evaluation studies:

### Scoping study

A scoping study will be carried out during the first months of the evaluation to determine the scope of the evaluation and engage with stakeholders.

### Study 1: Identifying goals, challenges and early lessons in implementation

Interviews at the outset with professionals who are the Generation Study’s “designers” and “early implementors”; shadowing of processes and pathways; and documentary analysis. Interviews towards the end of Generation Study recruitment with professionals including “designers” and “early implementers” and others who have become involved over time.

### Study 2: Impact, experiences and attitudes of parents

Survey and follow-up interviews with parents following receipt of gNBS results (Time 1) and 12 months later (Time 2) to assess acceptability, experience, attitudes and impact (positive and negative).

### Study 3: Gathering wider professional viewpoints about the Generation Study and gNBS implementation

Survey with staff delivering the Generation Study at early adopter sites and professionals from a range of relevant backgrounds across England.

### Study 4: Views of the rare disease community

Surveys and interviews with advocates from rare disease support groups to examine attitudes to gNBS including positive and negative impacts as well as unintended consequences on parents and the rare disease community more broadly.

### Study 5: Public views

Survey conducted with a market research company to capture views of with a diverse group of parents in terms of age, gender, ethnicity, socio-economic status and geographical location to evaluate the key findings around acceptability and attitudes towards gNBS with parents who have not been involved in the Generation Study.

### Study 6: Cost effectiveness evaluation

Assessment of the impact of gNBS on healthcare resource use and costs, quality of life outcomes and non-health-related outcomes, to inform the Genomics England health economics model.

### Study 7: Clinical utility assessment

Assessment of the clinical utility of gNBS for health-related outcomes by comparing hospital contact and mortality rates in various groups of children (‘condition suspected’ and false positives, population controls), making use of the longitudinal routinely collected healthcare data linked to the Generation Study and control groups primarily drawn from ECHILD.

## RESEARCH PLAN

### Scoping study

A scoping study will be carried out to determine the scope of the evaluation, identify and engage with key stakeholders and agree the final evaluation and dissemination plan with Genomics England. Discussions around the scope will include, prioritising the research questions set out by Genomics England, evaluation methods, timeline and dissemination plan. The scoping study will include meetings with key stakeholders and a review of relevant documents to understand the context of the Generation Study. We will also review the published and grey literature to identify any accessible study materials, (e.g. surveys and topic guides) being used in international evaluations of gNBS that could inform the development of our own study materials.

### Study 1: Identifying goals, challenges, and early lessons in implementation

#### Research questions

- Is the use of WGS as a tool for early diagnosis of rare, childhood-onset, actionable genetic conditions feasible and acceptable to professionals?
- What are the implementation processes being used to deliver the Generation Study and how do these vary across sites and over time?
- What are the effective approaches to implementation for delivery of gNBS including identified barriers and facilitators?
- What are professionals’ experiences of and attitudes towards delivering gNBS?
- What are professionals’ education and training needs to support delivery of gNBS?
- What factors impact access and uptake to the Generation Study?

#### Study design

The aim of this study is to understand the goals, challenges and early lessons of implementing gNBS in the Generation Study. This will include exploring the programme theory underpinning the Generation Study, assessing acceptability, feasibility, barriers and facilitators of implementation and impacts on staff time.

This will be done using five approaches;

- Qualitative interviews with 15-20 key stakeholders involved in designing, planning and discussing the Generation Study (“designers”); including professionals from organisations such as Genomics England, NHS England, clinicians from a range of specialties (both mainstream and genetics), policy makers and commissioners.
- Qualitative interviews with 5-6 professionals at each of the first 5-6 NHS Trusts undertaking recruitment to the Generation Study (“early implementers”); including clinicians from a range of specialties who are involved with recruitment and consent, returning results and study planning and management and staff education (e.g., midwives, genomics associates neonatal, paediatric and genetic specialists, researchers and data administrators and laboratory staff).
- Documentary analysis of relevant documents such as reports, policy documents, journal articles and meeting presentations.
- Shadowing of recruitment, consent and sampling processes at each of the first 5-6 NHS Trusts undertaking recruitment to the Generation Study.Follow-up interviews with a sub-set of the “designers” and “early implementers” of the Generation Study and interviews with other key professionals who have become involved over time.

#### Interview topic guides

Interview question guides have been developed with consideration for the existing literature, relevant theoretical frameworks (CFIR^3^ and NPT^4–6^) and the academic, clinical and PPI expertise of our research team. The topic guides will be continuously revised to address any new topics that emerge during the interviews. For the interviews with “designers”, the topic guide includes: Design of the Generation Study; Feasibility of implementing the Generation Study; Impact of the Generation Study; Barriers and facilitators; and Personal experience and reflections. For the interviews with “early implementors”, the topic guide includes: Design of the Generation Study; Acceptability of the Generation Study; Feasibility of implementing the Generation Study; Impact of the Generation Study; Barriers and facilitators; and Personal experience and reflections. For the follow-up interviews the topic guide includes: Acceptability of the Generation Study; Feasibility of implementing the Generation Study; Impact of the Generation Study; Barriers and facilitators; Changes to implementation strategies over time and Personal experience and reflections.

#### Data collection

Interviews will be carried out by phone, video call or face-to-face at the professional’s place of work. The interviews are expected to last approximately 45 to 60 minutes. Interviews will be conducted by a researcher experienced in qualitative interviews. Interviews will be digitally recorded, professionally transcribed verbatim and then anonymised.

Participant and non-participant observations will be carried out approximately 2-6 months after recruitment commences at each of the first 5-6 recruitment sites for the Generation Study. We will use a shadowing approach to observe and map how the Generation Study is delivered at each site including; how parents are identified and invited to take part in the Generation Study, length and content of consent discussions, processes for sample collection and dispatch, and which staff are involved.^15, 16^ Participant and non-participant observations will be carried out in the clinical areas where the discussions and tasks related to the Generation Study take place. Relevant staff meetings for observation will be identified by discussion with the Generation Study team at each site. These may include Generation Study team meetings, departmental meetings, multi-disciplinary team (MDT) meetings or educational events. The discussions between staff and parents will also be observed, this may include any initial discussions introducing the Generation Study to parents and taking consent to participate. A structured observation guide will be used to record all field notes to ensure consistency in the collection of data across researchers and sites. The field notes will record high-level summary information only. No identifying or confidential information about individuals will be recorded. This guide will be iteratively updated to explore themes emerging from the data.

To help us understand the reasons parents decline to take part in the Generation Study, a very brief ‘exit survey’ will be undertaken with those parents who decline when the Generation Study is discussed with them. This will be done by the health professional who is inviting the parents to take part in the Generation Study. Parents will be asked their reason(s) for declining to take part and if they choose to give a reason the health professional will record the response on a form that will comprise a short list of common reasons to decline and an open-ended question capture additional reasons.

Documentary evidence such as policy documents, journal articles and meeting presentations will be collated for analysis.

#### Sample size

We will conduct interviews with the “designers” and “early implementors” of the Generation Study until saturation is reached. Initially we anticipate conducting 15-20 interviews with “designers” and 25-30 interviews with the “early implementers” (5-6 from each site) based on previous similar research.^17, 18^ For the follow-up interviews, we anticipate conducting 25-30 interviews with a sub-set of “designers” and “early implementers” interviewed initially and other key professionals who have become involved in the Generation Study over time.

Observations will be conducted at each of the first 5-6 recruitment sites for the Generation Study. As part of the observations at each site, we will observe 4-5 discussions between staff and parents.

The exit survey will be conducted at 5-6 recruitment sites for the Generation Study. Professionals discussing the Generation Study with parents will be asked to complete the exit survey with any parents who decline within a 3 month period.

#### Recruitment

Interviews: The designers and early implementers of the Generation Study will be purposively sampled and invited to take part in a telephone or video interview. Relevant professionals will be identified by the research team with input from the Generation Study PIs at recruiting sites. “Snowball sampling” will also be used to ensure that key professionals not known to the team are invited. Professionals will be invited to take part in a semi-structured interview. Potential participants will be sent an invitation email along with a Participant Information Sheet describing the purpose of the interviews. The professionals will be asked to contact the research team via telephone or email if they are interested in participating in an interview. Prior to the start of the interview the participant will be asked if they have any questions, and it will be explained that the discussion will be recorded and transcribed but that no identifying information will be included on the transcript. Participants will then be asked to read and sign the consent form (or to give verbal consent (audio-recorded) for an interview conducted by telephone or video call). For the follow-up interviews, professionals who previously took part in an interview will be approached via email and invited to take part in a second interview. Purposive sampling will be used to identify other key professionals who have become involved in the Generation Study over time with the help of Genomics England and Generation Study PIs at the recruiting sites. Invitations and consent processes will be the same as the initial interviews.

Observations: Observations will be conducted at each of the first 5-6 recruitment sites for the Generation Study. We will adopt a ‘shadowing approach’ whereby a researcher shadows members of staff who are discussing and recruiting parents into the Generation Study. Observations will include the discussions professionals have with parents when first introducing the Generation Study, inviting them to take part and taking consent. These discussions may take place in the hospital or via phone or video call and this may differ between sites, depending on the individual processes developed for delivering the Generation Study at each site.

Prior to any observations taking place that involve both staff and parents, we will share a Participant Information Sheet with the professional and give them the opportunity to discuss the study with the researcher. The Participant Information Sheet will explain the purpose of the observation, the type of data to be collected (e.g. length of the discussion, topics discussed etc) and that the data collected will be anonymous. In the Participant Information Sheet and in the initial discussion with the researcher, it will be made clear that the health professional is required to gain verbal consent from parents for a researcher to observe the discussion. It will also be emphasised that the researchers should not have access to any confidential information about the parents prior to the parents giving consent for the researcher to observe the discussion. A guidance document to support health professionals to explain the purpose of the observation, the type of information collected and how to take verbal consent from the parents will be shared.

Observations of meetings: Before attending any specific staff meetings, the researcher will contact and seek permission to attend from the meeting Chair. A Participant Information Sheet for meeting attendees will be circulated to all attendees with the meeting papers; this will re-iterate the study details and will give an opportunity for any members of staff attending the meeting to opt out of the observation and not have their contributions to the meeting recorded in the field notes made by the researcher. The Participant Information Sheet will also be available at the beginning of the meeting. All field notes will be high level and no identifying or confidential details about individuals will be recorded.

Exit Survey: Professionals discussing the Generation Study with parents will be asked to complete the exit survey with any parents who decline to take part in the Generation Study within a 3 month period.

#### Data analysis

We will use both rapid qualitative data analysis and in-depth analysis.

##### Rapid qualitative data analysis

Analysis will be carried out in parallel with data collection and facilitated through interview notes and RREAL sheets; a working document where high level data are organised into categories.^19^ During the interviews and observations, the researchers will take notes that will be summarised and organised in the RREAL sheet immediately after each data collection episode. The RREAL sheets can then be used to summarise and share emerging findings on an ongoing basis.^19^ The RREAL sheets will also be used to identify topics that will need to be explored further using in-depth analysis.

##### In-depth analysis

Data from interviews, the exit survey and documents will be analysed using framework analysis.^20^ This is an approach that facilitates identification of key themes as well as commonalities and differences in the data through comparison within and across cases. The same framework will be applied to the analysis of both sets of interviews as well as the exit survey and documentary evidence to enable cross-referencing and comparisons across the data using a coding matrix in Excel.

### Study 2: Impact, experiences and attitudes of parents

#### Research questions

- Is the use of WGS as a tool for early diagnosis of rare, childhood-onset, actionable genetic conditions feasible and acceptable to parents?
- What are parents’ experience of joining the Generation Study?
- Did parents perceive that information about gNBS was effectively conveyed and that they were able to make an informed choice?
- What factors impact access and uptake to gNBS?
- What are parents’ information and support needs to facilitate decision making, receiving results and ongoing care and support networks?
- Can findings be returned effectively using the approach adopted for the study overall?
- What are parents’ experiences of receiving a ‘no condition suspected’ gNBS result?
- What are parents’ experiences of receiving a ‘condition suspected’ and a ‘confirmed diagnosis’ gNBS result, including the downstream pathways for parents receiving those results?
- What are the wider positive and negative impacts on families including parental mental health?
- What are parents’ experiences of receiving either a ‘no condition suspected’ or a positive gNBS result over the longer-term?

#### Study design

We will conduct a mixed-methods study including a Time 1 survey (n=500) and a sub-set of follow-up interviews (n=50) with parents following receipt of gNBS results to assess acceptability, experience, attitudes, quality of life and impact (positive and negative). The survey will be disseminated at least 3 months after parents receive gNBS results (including ‘no condition suspected’, ‘condition suspected’ and uncertain results). Three months was the timeframe considered acceptable by the PPI Advisory Group for parents who receive a “condition suspected” result from gNBS. As such, we will send the Time 1 survey to all potential participants at this same timepoint. Parents who complete a Time 1 survey will be invited to complete a Time 2 survey around 12 months after completing the Time 1 survey. A sub-set of Time 2 survey respondents will be invited to take part in a Time 2 interview.

#### Survey content

The Time 1 and Time 2 surveys include a mix of bespoke questions and validated measures and were informed by our study and PPI advisory groups. The Time 1 survey will follow on from the baseline survey conducted by Genomics England to explore parents’ experience of the consent process, which will include, amongst other questions, the EQ-5D-5L^21^ to assess parental quality of life, the GAD-7 anxiety measure^22^ and the SURE measure of decisional conflict.^23^ }

For the Time 1 survey, to measure quality of life and non-health outcomes we are including the following measures: <u>parental quality of life including</u> (the EQ-5D-5L^21^ and PedsQL Family Impact survey^24^ [for parents who received a true positive (condition confirmed) result only]); <u>child quality of</u> <u>life</u> (EQ-TIPS^25^ which is a proxy-reported instrument designed for infants and toddlers); anxiety (GAD-7^22^) and <u>non-health outcomes</u> including parental personal utility (GENE-U^26^ for parents who received a true positive (condition confirmed) result only]); parent-child bonding (Mother Infant Bonding scale^27^); parents’ perception of child’s vulnerability (Vulnerable Baby Scale^28^); decisional regret (Decisional Regret scale^29^); family experience of joining the programme including satisfaction with receipt of results; signposting to support groups and impact on wider family e.g. key decision-makers, cascade testing. We will also ask parents to recall any out of pocket costs incurred from consent to results receipt.

The Time 2 survey includes the validated measures that were included in the Time 1 survey, to allow for comparisons over time, and several bespoke questions. The bespoke questions relate to participants’ subsequent pregnancies and intentions to take part again and recommendations to other parents.

#### Interview topic guides

Time 1 interviews will be conducted with a subset of Time 1 survey responders who have indicated on the survey that they are willing to take part in an interview, expected to last 30-45 minutes. We will purposively select participants to ensure a range in terms of participant characteristics and gNBS results. The interview topic-guide was co-designed with the study and PPI advisory groups. Interview topics include: acceptability of gNBS, perceived benefits and concerns, experience of joining the Generation Study (satisfaction with the consent process, information and support needs, timing of approach, turnaround time, receiving results, ongoing care and support networks), and positive and negative impact of receiving gNBS results including psychological impact, any unanticipated outcomes and perceived utility e.g., for reproductive decision-making, other family members.

Time 2 interviews will be conducted with a subset of Time 2 survey responders who have indicated their willingness to take part in an interview on the survey. We will purposively select participants to ensure a range in terms of participant characteristics and gNBS results. The Time 2 interview topic guide builds on the Time 1 topic guide and includes: acceptability of gNBS, perceived benefits and concerns, experience of taking part in the Generation Study (ongoing information and support needs), experience of NHS healthcare after results returned, any regrets about taking part and positive and negative impact of receiving gNBS results including psychological impact, any unanticipated outcomes and perceived utility e.g., for reproductive decision-making, cascade testing for other family members.

#### Data collection

At Time 1, potential participants will be identified by the research team at Genomics England and sent an email or letter by post inviting them to complete the survey. At Time 2, an invitation to complete the surveys will be sent via email or letter (if previously preferred) to participants who completed a Time 1 survey. Surveys can either be completed on paper and sent back using a freepost envelope, by phone with a researcher or online using REDCap (a link and QR code will be included in the invitation). The survey will take 20-30 minutes to complete.

Interviews will be carried out by phone or video call. The interviews are expected to last approximately 45 to 60 minutes. Interviews will be conducted by a researcher experienced in qualitative interviews. Interviews will be digitally recorded, professionally transcribed verbatim and then anonymised. If participants prefer to not have the interview recorded, then the interviewer will take written notes during the interview.

#### Sample size

For the Time 1 survey, we have estimated a sample size of 500 completed surveys. This is based on our previous survey study where we recruited 504 participants taking part in the 100,000 Genomes Project across 6 sites in 15 months.^30^ A sample size of 500 would allow for a difference in the mean score of 5 between those receiving a ‘condition suspected’ and those receiving a ‘no condition suspected’ gNBS result on the decisional regret scale (at 80% power, α = .05). We also aim to include a subset of responders who receive a ‘false positive’, ‘false negative’ or ‘uncertain’ result. All participants who complete a survey at Time 1 will be invited to complete a survey at Time 2. Based on similar studies, we anticipate a 60% response rate to give a sample size of 300.

We anticipate interviewing ∼50 parents at both Time 1 and Time 2 so that we can include parents from a broad range of participant characteristics.

#### Sampling and recruitment

Time 1 survey participants will be a sub-set of participants recruited into the Generation Study. We will recruit participants across demographic groups e.g. ethnicity, education, health literacy, gender using a stratified random sampling approach to explore the role of health inequalities. Genomics England will send an invitation to complete the survey by email or letter to the potential participants. The email will include a Participant Information Sheet describing the survey and interview study and a link to complete the survey online. Potential participants will be able to request a paper copy of the survey that can be returned by reply-paid post. Recruitment will continue until the minimum number of completed surveys have been collected. At Time 2, an invitation to complete the surveys will be sent via email or letter (if previously preferred) by Genomics England to participants who completed a Time 1 survey. All participants who complete a survey will be sent a gift voucher in appreciation of their time.

A subset of survey responders will be purposively sampled to take part in an interview at Time 1 and Time 2, aiming for a range in terms of gender, age, ethnicity, education, health literacy, place of enrolment, parity, type of gNBS results received. Participants who take part in an interview will be sent a gift voucher in appreciation of their time.

The contact details of the research team will be included in the first page of the Time 1 and Time 2 survey, so that potential participants can contact the research team with any queries. If participants do feel distressed by the survey or interview, the participant information sheet contains several alternatives for seeking advice or support including, a counsellor from the organisation Rareminds (https://www.rareminds.org/) who provide mental health support for the rare disease community and are independent of the Generation Study, their regional clinical genomics service, their general practitioner (GP) or their Trust Chaplains.

#### Data analysis

The Time 1 and Time 2 surveys will be analysed using both descriptive and inferential statistics to gain a nuanced understanding of acceptability and uptake amongst different population groups. For the analysis of the interviews will use rapid qualitative data analysis so that results can be fed back quickly, which will be followed by in-depth analysis.

##### Rapid qualitative data analysis

As described for Study 1, rapid qualitative data analysis will be carried out in parallel with data collection and facilitated through interview notes and RREAL sheets.^19^ The RREAL sheets will be used to summarise and share emerging findings on an ongoing basis.^19^ The RREAL sheets will also be used to identify topics that will need to be explored further using in-depth analysis.

##### In-depth analysis

Data from the interviews will be analysed using thematic analysis.^31^ Analysis will involve an iterative process where data are coded, compared, contrasted and refined to generate themes. Analysis will be conducted by at least two researchers to provide rigour. Coding will be informed by the topic guide (deductive analysis) and may also include new topics or unexpected findings (inductive analysis).^32^ Data analysis will be facilitated by using a coding matrix in Excel and, as needed, NVivo version 13 (QSR International, Pty Ltd).

#### Linkage to the Generation Study data

Each parent invited to complete a survey will be assigned a unique code by Genomics England to facilitate tracking and linkage. The survey data will be stored on and analysed using the UCL Data Safe Haven. This data-set will also be shared securely with Genomics England and the unique code will be used to link an individual’s survey data with the Generation Study data including diagnostic outcomes data and longitudinal secondary datasets within the secure environment of the National Genomic Research Library (NGRL). Only relevant members of the Generation Study and Evaluation Study team’s will be able to access these linked data sets, which will be stored within a ring-fenced area of Genomics England’s secure NGRL, which will not be accessible to other researchers. Any practices in relation to the NGRL will be carried out in line with Genomics England’s NGRL approved protocol.^33^

### Study 3: Gathering wider professional viewpoints about the Generation Study and gNBS implementation

#### Research questions

- Is the use of gNBS as a tool for early diagnosis of rare, childhood-onset, actionable genetic conditions feasible and acceptable to the wider workforce?
- What are wider workforces’ experiences and attitudes to the use of gNBS as a tool for early diagnosis of rare, childhood-onset, actionable genetic conditions?
- What are the downstream implications of offering gNBS including impacts on the wider clinical workforce?

#### Study design

The aim of this study is to obtain a broader understanding of professional views and experiences of offering gNBS in the Generation Study and gather wider opinions around introducing gNBS into clinical practice as part of routine care. To do this we will develop an anonymous cross-sectional survey that will be circulated after the Generation study has been scaled-up for implementation at multiple sites. Potential participants will include: A. professionals delivering the Generation Study at all NHS Trusts that are actively recruiting, B. professionals from the NHS Trusts where babies with a condition suspected result from gNBS have been referred for return of results and clinical care, and C. professionals from a range of relevant backgrounds from across England who are not directly involved with delivery of the Generation Study.

#### Survey content

^3435^Survey development will be informed by the existing literature, findings from the qualitative interviews with professionals in Study 1 and the expertise of our advisory groups. The survey will include a mix of validated measures and bespoke questions. The validated measures address the concepts of attitude^36^ and acceptability.^35^ Other questions include: demographics, awareness and knowledge of genomics, blood spot newborn screening and the Generation Study, perceived benefits and concerns about genomic newborn screening and the Generation Study, challenges for delivering gNBS in routine care, expected workforce implications, potential impacts on healthcare systems, education and training needs and overall views. The survey will also capture differences or similarities in processes used by Generation Study recruitment sites for recruitment, supporting parents from diverse backgrounds, use of interpreters and reasons to decline. Responses will be anonymous. Survey development has included several stages of review and revision with feedback obtained from the wider research team, members of our Study Advisory Group and our PPI Advisory Group and professionals from Genomics England working on the Generation Study. The survey has also been reviewed by professionals from different backgrounds to ensure consistency and clarity.

#### Data collection

The survey will be hosted online using REDCap and will be made available for 3 months. Completion of the survey is expected to take approximately 20-25 minutes per participant.

#### Sample size

Participants will be purposefully sampled to ensure there is maximum variation professional background and geographical location. A sample size of 200, including 5-10 participants from each Generation Study recruitment site and 10-15 from each of the seven GLH/GMSA regions in England has been estimated on the basis of our previous research experience as being sufficient to provide a depth and breadth of opinions and attitudes. Due to the nature of the survey, it is not proposed that this sample will be statistically representative of the population groups consulted.

#### Sampling and recruitment

Recruitment will involve two approaches. 1. The Generation Study PI(s) at each recruitment site will be asked to provide a list of potential participants that will include professionals involved in recruitment and consent for the Generation Study, professionals from the NHS Trusts where babies with a positive result from gNBS have been referred for return of results and clinical care and professionals involved in local care pathways relevant to newborn screening. The research team will circulate an invitation email with a link to the online survey to the potential participants. Three reminder emails will be sent. 2. A study flyer with a link to the survey will be circulated through social media and email lists of professional bodies such as the Royal College of Midwives, the Royal College of Paediatrics and Child Health, the Royal College of Nursing and the British Society for Genetic Medicine.

#### Data analysis

Quantitative data from the survey will be analysed using descriptive statistics and open-ended survey questions will be analysed using thematic analysis.^31^

### Study 4: Views of the rare disease community

#### Research questions

- How prepared are patient organisations to support families taking part in gNBS?
- What impact do patient organisations think gNBS will have on the families they support now and in the future?

#### Study design

Many rare disease patient advocacy organisations provide direct support to affected individuals e.g. through helplines, as well as working to influence the services and support available through statutory services. Many are small organisations with few staff, sometimes working as volunteers. To examine views on gNBS from the perspective of these organisations and the rare disease community more broadly, we will conduct surveys and interviews with advocates from rare disease support groups.

#### Survey content

An online survey will be co-produced with our PPI Advisory Group and representatives of a small number of patient advocacy organisations. Questions to be explored will include: Do rare disease advocacy organisations have concerns about the Generation Study and gNBS generally in terms of its impact on them or the people they support? Were they able to take part in consultations run by Genomics England before the study was launched, and what are their views of these consultations? Are there changes in demand for support and are they prepared (greater volume/new situations e.g. pre-symptomatic diagnoses)? How are parents finding them (e.g. signposted from clinical services)? What do they see as benefits for their community? Do they foresee any unintended consequences?

#### Interview topic guides

Interviews will be conducted with a subset of survey responders. The interview topic-guide will be co-produced with the PPI Advisory Group. Interview topics reflect the survey content, providing an opportunity to discuss these issues in more depth.

#### Data collection

The survey will be hosted online through REDCap and will be made available for 6 weeks. Completion of the survey is expected to take approximately 20-25 minutes per participant. Interviews will be carried out by phone or video call. The interviews are expected to last approximately 45 to 60 minutes. Interviews will be conducted by a researcher experienced in qualitative interviews. Interviews will be digitally recorded, professionally transcribed verbatim and then anonymised. If participants prefer not to have the interview recorded then the interviewer will take written notes during the interview.

#### Sample size

There are over 200 organisations in the UK who provide support to parents and families impacted by rare diseases. Based on previous surveys with these groups, we anticipate around 50% of the organisations will respond, giving us a sample size of 100 (Amy Hunter, personal communication), which will be sufficient to provide a depth and breadth of opinions and attitudes. Follow-up qualitative interviews will be arranged with ∼10-20 representatives of rare disease organisations (10%).

#### Sampling and recruitment

An invitation email that will include the Participant Information Sheet and a link to the online survey will be circulated by the research team at Genetic Alliance UK to the ∼230 member organisations of Genetic Alliance UK, and the organisational members of The Neurological Alliance who encompass rare conditions. Three reminder emails will be sent during the six weeks the survey will be open.

At the end of the survey respondents will be asked if they would be interested in taking part in a follow-up interview. Survey participants invited to take part in a follow-up interview will be purposively sampled to ensure we gather more in-depth views from organisations from a range of sizes and representing a range of different types of conditions.

#### Data analysis

Descriptive statistics will be generated from the survey data. Open text responses to the survey and qualitative interview transcripts will be analysed using thematic analysis.^31^

### Study 5: Public views

#### Research questions

- What are parents from the general public (i.e. those who have not been invited to take part in the Generation Study) attitudes towards gNBS?
- Do parents from different ethnic or educational backgrounds differ on their views of gNBS?
- Do parents from the general public view gNBS as acceptable?

#### Study design

We will conduct a survey with parents from the public who have given birth in the previous two years and who have <u>not</u> been invited to take part in the Generation Study. The survey will ascertain the views of parents towards gNBS and in particular to examine whether the acceptability and attitudes identified through our survey and interviews with parents in Study 2 are shared by the public. The survey will be conducted with the market research company Dynata (https://www.dynata.com/), who we have worked with previously.^37^ A service agreement will be in place between Dynata and UCLC.

#### Survey content

An online survey will assess acceptability, views and attitudes towards gNBS. Survey development will be informed by the findings from the surveys and interviews with parents taking part in the Generation study (Study 2) and the expertise of our Study and PPI advisory groups.

#### Data collection

We will conduct an online survey that will be set up on REDCap through the market research company Dynata.

#### Sample size

As the survey will be conducted through a market research company, we will request quotas for specific population groups, in particular, those whom we may not have reached through our parent survey. We will recruit 200-250 survey responders to enable sufficient numbers to compare across groups. An a priori power analysis was conducted to determine the minimum sample size required to test the study hypothesis. The required sample size to achieve 80% power for detecting a medium effect, at a significance criterion of α = .05 was n = 216 for tests of comparisons across sub-groups.

#### Recruitment

Recruitment will be done by the market research company Dynata. An invitation to complete an online survey will be circulated by Dynata to women and men (over the age of 18) who have had a child in the last two years. This will ensure that we capture the views of parents who have recently had a child and have been through antenatal and neonatal services.

In the first instance, Dynata, will send the survey link to a small group of potential participants as a pilot. The aim of the pilot will be to test out the recruitment method and to ensure the survey is completed as intended. We will aim to collect 20 completed surveys in the pilot study. Following the pilot, the survey will be sent to a larger number of potential participants and recruitment will continue until the target sample size is reached across all requested population groups. Dynata will send an invitation to participate with a link to complete the survey by email or potential participants will access Dynata’s system themselves to look for surveys. Participants who complete the survey will be paid Dynata’s standard E-reward payment that is equivalent to £1.00 for a 20 minute survey.

The contact details of the research team will be included in the first page of the survey, so that potential participants can contact the research team with any queries. To make sure parents also have a point of contact for support if the survey content raises any worries or questions, at the end of the survey we have provided advice for seeking support from their GP or a counsellor from the organisation rareminds (https://www.rareminds.org/) who provide mental health support for the rare disease community and are independent of the Generation Study.

#### Data analysis

Survey data will be analysed using descriptive and inferential statistics.

### Study 6: Cost effectiveness evaluation

#### Research questions

- What is the impact on secondary healthcare resource use including estimates of end-to-end costs – from recruitment, through sample processing, sequencing and interpretation through to clinical care?
- What are the positive and negative impacts on health-related outcomes including: morbidity, mortality and quality of life?
- What is the impact on non-health-related outcomes including: perceived personal utility and psychosocial wellbeing?

#### Study design

We will evaluate the impact of gNBS on healthcare resource use and costs, quality of life outcomes and non-health-related outcomes. Parameter estimates will be generated to support Genomics England with the development of an economic model to answer the primary cost-effectiveness research question. Specifically, we will:

1. Estimate the costs associated with sequencing newborns in the Generation Study, and the costs of implementation and scaling up;
2. Estimate the impact of gNBS on healthcare resource use and associated costs, and on out of pocket costs for children and their families, compared to children who do not undergo sequencing;
3. Estimate the impact of gNBS on morbidity and quality of life outcomes in children, and quality of life outcomes in parents, compared to children who do not undergo sequencing and their parents;
4. Estimate the impact of gNBS on non-health-outcomes for parents, compared to parents of children who do not undergo sequencing.

#### Data collection and sampling

A microcosting study will be conducted alongside Study 1 to estimate the costs associated with sequencing children, from sample collection to return of results. The microcosting study will capture resource utilisation associated with delivering gNBS as a screening service during the initial period of the Generation Study, and will then be repeated later in the study. The purpose of the two microcosting analyses is to identify changes in resource use over time and assess efficiencies associated with the study’s ongoing learning and increased recruitment. Current screening costs will be extracted from the literature.^38, 39^ Data on the costs of implementation and scaling up will be collected in the interviews (Study 1) and cross-sectional survey with professionals (Study 3) and combined with data on likely uptake (Study 2).

The analysis of secondary care resource use for children undergoing gNBS will use the linked Hospital Episode Statistics data made available by Genomics England, combined with NHS Reference Costs. Secondary care resource use and costs for participants in the 100,000 Genomes Project, and for ‘no condition suspected’ participants in the Generation Study, will be used as controls. Data on out of pocket costs accrued by participants in the Generation Study (e.g. over-the-counter medications, caregiver time) will be collected via Study 2 of the Impact Evaluation, at both the Time 1 survey (3 months after parents receive gNBS results) and Time 2 survey (12 months after completing the Time 1 survey). Comparator costs will be extracted from the literature.^40, 41^ Analysis of morbidity and mortality outcomes will use data made available in the Generation Study for newborns undergoing sequencing, and for age matched children with comparable diagnoses who have not undergone sequencing.

Data on quality of life outcomes in newborns will be collected using proxy-completed age-specific instruments (EQ-TIPS^25^), administered in the parent survey at both Time 1 and Time 2 (Study 2). Baseline values are available in the literature^42^ and from the instrument developers. Data on the quality of life of parents will be collected using multiple instruments (EQ-5D-5L,^21^ GAD-7^22^, PedsQL Family Impact Module^43^), via the parent surveys in Study-2. The baseline values of the EQ-5D-5L and GAD-7 will be measured through consent survey distributed by Genomics England whereas the baseline values of the PedsQL Family Impact Module will be derived from literature sources. Quality of life data for children not undergoing sequencing, and their parents, will be estimated using established methods.^38^ Non-health outcomes will be measured using multiple instruments (GENE-U^26^, Vulnerable Baby Scale^28^, Mother to Infant Bonding Scale^27^, Decision Regret Scale^29^). Data will be collected in the parent survey (Study 2) and supplemented with data from other newborn sequencing studies applying these instruments (e.g. BabySeq 2), via established collaborations.

#### Data analysis

For all parameters, descriptive statistics will be presented (e.g. mean values, standard deviations, confidence intervals). Summary statistics for cost parameters will be expressed across specific time periods (e.g. per month/year) to align with the economic model. Results for quality of life instruments will be presented at the domain level and, where value sets exist, as utility scores. Summary statistics will be presented for predefined subgroups (e.g. by sequencing result, by condition). Differences will be calculated between parameter values at baseline and after the return of results. If required for the economic model, we will use quasi-experimental approaches (e.g. difference-in-difference) to further quantify differences in outcomes between sequenced and non-sequenced children. Threshold analyses will explore the impact of key assumptions. Missing data will be quantified as required, and analyses adjusted using appropriate methods (e.g. multiple imputation).

### Study 7: Clinical utility assessment

#### Research questions

- What are the positive and negative impacts on health-related outcomes (morbidity, mortality)?
- What is the prevalence of the conditions looked for in the newborn population?
- How many are ‘confirmed diagnosis’ and how many would be expected if it were adopted nationally?
- What is the level of uptake of the programme and what would the level of uptake of genomic newborn screening be if it were adopted nationally?

#### Study design

We will use a series of cohort studies to assess the impact of gNBS on health-related outcomes. Our objectives are to:

1. Estimate hospital contact & mortality rates among children who are confirmed to have a rare condition via gNBS, compared to children with similar conditions who were diagnosed through routine clinical practice
2. Estimate hospital contact & and mortality rates among children who are ‘false positive’, compared to a) children in the Generation Study who receive a ‘no condition suspected’ result, and b) the general population of children in England
3. Estimate the impact of introducing the programme in England on health service use and mortality, taking into account differential selection into the Generation Study

#### Data sources

We will utilise longitudinal healthcare data that are being linked to the Generation Study (including HES, ONS mortality data, the NNRD). We will work with Genomics England to determine the optimal datasets for control groups, however we propose to draw comparator data from the Education and Health Education and Child Health Insights from Linked Data (ECHILD). (https://www.ucl.ac.uk/child-health/research/population-policy-and-practice-research-and-teaching-department/cenb-clinical-20) ECHILD links administrative health data, including HES, to education data for all individuals born in England since 1997. A mother-baby link is being incorporated into ECHILD, allowing future studies of the health of mothers and siblings of children with rare conditions.

#### Data analysis

The primary outcome will be emergency hospital contact (accident and emergency [A&E] attendances and admission rates, derived from HES. Our secondary outcomes will be NICU admission rates (from NNRD), planned hospital admission rates and mortality. Our primary focus is on secondary care use; we expect a small number of deaths in the recruited cohort. Given the start date of recruitment, the oldest children in the Generation Study will be over 12 months of age at the start of the study. Early development and education outcomes will therefore be out of scope, however we will develop analysis approaches and code to be applied in future natural history studies using Community Services Dataset and National Pupil Data (part of ECHILD).

Objective 1: We will use linked Generation Study-HES-mortality data from the NGRL to calculate A&E attendance, emergency hospital admission and mortality rates among true positive children. We expect to group children with similar conditions for analyses; useful groups will be agreed with the Genomics England Team. We will use the linked Generation Study-HES data for true positive children to define these conditions in HES using clinical code lists. We will apply these definitions in ECHILD and estimate prevalence of these conditions in general population, hospital use and mortality among children with similar conditions diagnosed via routine practice. This approach can be extended to examine early development (via CSDS) and National Pupil data.

Objective 2: We will estimate hospital admission and mortality rates among ‘false positive’ and ‘no condition suspected’ children using the linked Generation Study-HES-mortality data. Several population control groups can be derived using ECHILD, including all children born in England during particular time periods, children meeting inclusion criteria born in participating trusts during non-recruiting periods, or children meeting inclusion criteria born during recruiting periods but in non-participating trusts – to be agreed with Genomics England. For both objective 1 and 2, we will use generalised linear models and/or survival analyses to examine impact of gNBS on hospital admission and mortality rates.

Objective 3: We will use results from objective 2 and 3 applied to ECHILD data to estimate the number of hospital admissions and deaths prevented by gNBS across England. We will also use ECHILD data to estimate level of gNBS uptake and screen positives if the programme was applied nationally. We will use inverse probability weighting and similar methods to account for likely under-representation of some groups of children consented to the Generation Study (such as children from minority ethnic groups and more deprived families), and the exclusion of premature babies, to examine gNBS’s impact on health inequalities.

#### Integration of findings

Research findings from all studies will be integrated using a mixed-methods coding matrix that will be linked to each research question. Drawing together the findings of the individual studies will generate a holistic picture of the outcomes of our Process and Impact Evaluation.

## PROJECT MANAGEMENT

### Team working

The full research team will meet at fortnightly ‘check-in’ meetings (videoconference) to monitor overall progress against the project plan. At these meetings the lead research for each study will be asked to feedback progress against their deliverable(s). Any potential issues or delays will be discussed amongst the team, including whether wider discussion with the advisory teams is necessary. Smaller working groups will be established to collaborate on individual studies. UCL Consultants Ltd (UCLC) will provide project management, financial monitoring and risk management support throughout the evaluation. UCLC will also take responsibility for contracts and account management.

### Data sharing

Non-sensitive data such as data collection documents and study reports will be shared through the Office Teams platform which all members of the research team will have access to. De-identified research data such as anonymised survey data will be stored on the password protected UCL network. Personal data such as contact details DoB, ethnicity etc as well as survey responses, recordings of interviews and non-anonymised transcripts will be stored on the Data Safe Haven. Sensitive data will only be accessible to the research team through the Data Safe Haven. Any paper copies of surveys will be kept in a secure UCL office.

### Collaboration with Genomics England

We will share formative feedback at a virtual meeting with Genomics England every month to share our findings as they emerge. Findings will be shared via slide presentations with an agreed template. Formative feedback will include (1) sharing our evolving understanding of the programme theory and suggested refinements; (2) providing ‘real-time’ insights on the implementation of the Generation Study; (3) presenting staff views on the processes of implementation, (4) sharing insights on the experiences of parents from surveys and interviews and (5) discussing interpretation of findings. Formative reporting will also include interim reports delivered in January 2025, May 2025, December 2025 and December 2026 and the final report delivered in March 2028.

## ETHICAL AND REGULATORY COMPLIANCE

This study will be conducted in line with the ethical framework set out by the NHS Health Research Authority and according to Good Clinical Practice (GCP) principles. The study design includes measures to safeguard the wellbeing and dignity of participants and all staff and procedures will comply with the Data Protection Act 1998 and the EU General Data Protection Regulation. Recruitment will only take place once Research Ethics Committee (REC) and Health Regulatory Authority (HRA) approvals are in place. The Generation Study and the Process and Impact Evaluation have been approved by the HRA and the East of England – Cambridge Central NHS REC (23/EE/0044).

## RISK MANAGEMENT

UCLC has incorporated a risk management strategy and policy as part of its internal control and corporate governance arrangements. UCLC applies best practice in the identification, evaluation and cost effective control of risks to ensure that they are eliminated or mitigated. Risk considerations will be assessed throughout the project life cycle. UCLC will work with the research team and Genomics England to establish a Risk Register and appropriate escalation and risk management approaches at the project outset. The Risk Register will be a live document on a Teams site in which the project team can review the existing risks / add items at any time. The project manager from UCLC will be responsible for maintaining and updating the Risk Register, however, specific risks will be delegated to the relevant research team members. Updates to the Risk Register will be an agenda item at the research team’s fortnightly check-in meetings so that any identified risks can be discussed and addressed rapidly. Any potential issues or delays will be discussed amongst the team, including whether wider discussion with the advisory team or Genomics England is necessary.

## DISSEMINATION

Our strategy for engagement, formative feedback and dissemination includes:

- Peer reviewed scientific publications.
- Presentations to scientific meetings, nationally and internationally.
- Feedback of findings to professionals from a range of backgrounds.
- Plain language summaries of findings, written with the help of the PPI Advisory Group, will be disseminated to parent and patient networks via meetings, newsletters and social media.

Authorship eligibility guidelines and any intended use of professional writers Eligibility for authorship will follow the guidelines set out by the International Committee of Medical Journal Editors (ICMJE) (https://www.icmje.org/recommendations/browse/roles-and-responsibilities/defining-the-role-of-authors-and-contributors.html). No professional writers will be used.

## APPENDICIES

1. Questions in the Genomic England ITT not covered in the Process and Impact Evaluation that will be covered internally by Genomics England and/or through other collaborations

### Feasibility and Acceptability

Whether approaches to implementation could scale

Can samples be taken consistently in a busy newborn setting and with sufficient quality to support WGS and analysis?

Can a sufficiently rapid end to end turnaround time be achieved from sample to issue of (positive and negative) screening results to families (including confirmatory testing) to inform clinical care?

Can findings be returned safely and effectively using the approach adopted for the study overall?

Are the specific clinical pathways established for the disorders included in the study being followed?

Can we sustainably collect data on important outcomes for babies and families, and is the process of collecting this outcome sustainable to families?

### Impact

What is the impact of the programme on families, stakeholders and the wider system?

What is the impact on the wider family and society including: family member case identification, reproductive choices and patient and public values and preferences?

### Test performance and clinical utility

What is the clinical utility of genomic newborn screening as judged by the number of apparent true positive screening diagnoses identified?

What proportion of apparent false positive and false negative findings are there in the study according to each condition’s working case definition?

What proportion of babies have uncertain status following orthogonal testing according to each condition’s working case definition?

What age are looked for conditions clinically diagnosed and treatment started with genomic newborn screening as compared to standard of care alone?

What are the demographic factors that determine outcome e.g., percentage apparent true positive findings?

## Competing Interest Statement

James Buchanan has received travel support from Illumina and consulting fees from Genomics England.

Martin Vu has received a PhD scholarship from Illumina under the University of Melbourne/Illumina partnership.

Amy Hunter and Jennifer Jones are employees of Genetic Alliance UK. Genetic Alliance UK runs Rare Disease UK – a campaign for people with rare diseases and all who support them. Rare Disease UK receives financial support from a range of companies developing therapeutics for rare conditions.

All other authors have declared no competing interest.

## Funding Statement

This manuscript presents independent research commissioned by Genomics England. The views expressed are those of the authors and not necessarily those of Genomics England, the NHS or the UK Department of Health.

## Data Availability Statement

No data are associated with this manuscript as it is a study protocol.

## Notes

### Author Declarations

The Health Research Authority (HRA) and the East of England Cambridge Central NHS Research Ethics Committee gave ethical approval for this work (23/EE/0044).

### Summary of Updates

The protocol has been updated to include a longitudinal assessment of parent and professional experiences.

## REFERENCES

1. Andrew S, Halcomb EJ. Mixed methods research is an effective method of enquiry for community health research. Contemp Nurse 2006;23:145–53. 10.5555/conu.2006.23.2.145

2. Allemang B, Sitter K, Dimitropoulos G. Pragmatism as a paradigm for patient-oriented research. Health Expect 2022;25:38–47. 10.1111/hex.13384

3. Damschroder LJ, Reardon CM, Widerquist MAO, Lowery J. The updated Consolidated Framework for Implementation Research based on user feedback. Implementation Science 2022;17:75. 10.1186/s13012-022-01245-0

4. May C. Towards a general theory of implementation. Implementation Science 2013;8:18. 10.1186/1748-5908-8-18

5. May C, Finch T, Mair F, Ballini L, Dowrick C, Eccles M, et al. Understanding the implementation of complex interventions in health care: the normalization process model. BMC Health Serv Res 2007;7:148. 10.1186/1472-6963-7-148

6. McEvoy R, Ballini L, Maltoni S, O’Donnell CA, Mair FS, MacFarlane A. A qualitative systematic review of studies using the normalization process theory to research implementation processes. Implementation Science 2014;9:2. 10.1186/1748-5908-9-2

7. Orlando LA, Sperber NR, Voils C, Nichols M, Myers RA, Wu RR, et al. Developing a common framework for evaluating the implementation of genomic medicine interventions in clinical care: the IGNITE Network’s Common Measures Working Group. Genet Med 2018;20:655–63. 10.1038/gim.2017.144

8. Best S, Brown H, Lunke S, Patel C, Pinner J, Barnett CP, et al. Learning from scaling up ultra-rapid genomic testing for critically ill children to a national level. NPJ Genom Med 2021;6:5. 10.1038/s41525-020-00168-3

9. Allen CG, Sterba K, Norman S, Jackson A, Hunt KJ, McMahon L, Judge DP. Use of a multi-phased approach to identify and address facilitators and barriers to the implementation of a population-wide genomic screening program. Implement Sci Commun 2023;4:122. 10.1186/s43058-023-00500-9

10. Kansal A, Quinlan C, Stark Z, Kerr PG, Mallett AJ, Lakshmanan C, et al. Theory Designed Strategies to Support Implementation of Genomics in Nephrology. Genes (Basel) 2022;13. 10.3390/genes13101919

11. Keith RE, Crosson JC, O’Malley AS, Cromp D, Taylor EF. Using the Consolidated Framework for Implementation Research (CFIR) to produce actionable findings: a rapid-cycle evaluation approach to improving implementation. Implement Sci 2017;12:15. 10.1186/s13012-017-0550-7

12. Cresswell JW, Plano Clark VL. Designing and Conducting Mixed Methods. Research. Thousand Oaks, CA: Sage Publications, Inc.; 2011.

13. Vindrola-Padros C, Brage E, Johnson GA. Rapid, Responsive, and Relevant?: A Systematic Review of Rapid Evaluations in Health Care. American Journal of Evaluation 2021;42:13–27. 10.1177/1098214019886914

14. Vindrola-Padros C. Rapid Ethnographies: A Practical Guide. Cambridge: Cambridge University Press; 2021.

15. Martin S, Clark SE, Gerrand C, Gilchrist K, Lawal M, Maio L, et al. Patients’ Experiences of a Sarcoma Diagnosis: A Process Mapping Exercise of Diagnostic Pathways. Cancers (Basel) 2023;15:3946. 10.3390/cancers15153946

16. Bedwell GJ, Dias P, Hahnle L, Anaeli A, Baker T, Beane A, et al. Barriers to Quality Perioperative Care Delivery in Low- and Middle-Income Countries: A Qualitative Rapid Appraisal Study. Anesth Analg 2022;135:1217–32. 10.1213/ane.0000000000006113

17. Sanderson SC, Hill M, Patch C, Searle B, Lewis C, Chitty LS. Delivering genome sequencing in clinical practice: an interview study with healthcare professionals involved in the 100 000 Genomes Project. BMJ Open 2019;9:e029699. 10.1136/bmjopen-2019-029699

18. Clark CCA, Boardman FK. Expanding the notion of “benefit”: comparing public, parent, and professional attitudes towards whole genome sequencing in newborns. New Genetics and Society 2022;41:96–115. 10.1080/14636778.2022.2091533

19. Vindrola-Padros C, Chisnall G, Polanco N, San Juan NV. Iterative cycles in qualitative research: Introducing the RREAL Sheet as an innovative process. OSF Preprints, June 25 doi:1031219/osfio/9dp2w 2022. https://doi.org/

20. Gale NK, Heath G, Cameron E, Rashid S, Redwood S. Using the framework method for the analysis of qualitative data in multi-disciplinary health research. BMC Med Res Methodol 2013;13:117. 10.1186/1471-2288-13-117

21. Devlin N, Pickard S, Busschbach J. The Development of the EQ-5D-5L and its Value Sets. In: Devlin N, Roudijk B, Ludwig K, editors. Value Sets for EQ-5D-5L: A Compendium, Comparative Review & User Guide. Cham (CH): Springer Copyright 2022, The Author(s). 2022. p. 1–12.

22. Spitzer RL, Kroenke K, Williams JB, Lowe B. A brief measure for assessing generalized anxiety disorder: the GAD-7. Arch Intern Med 2006;166:1092–7. 10.1001/archinte.166.10.1092

23. Légaré F, Kearing S, Clay K, Gagnon S, D’Amours D, Rousseau M, O’Connor A. Are you SURE?: Assessing patient decisional conflict with a 4-item screening test. Can Fam Physician 2010;56:e308–14. https://doi.org/

24. Varni JW, Sherman SA, Burwinkle TM, Dickinson PE, Dixon P. The PedsQL Family Impact Module: preliminary reliability and validity. Health Qual Life Outcomes 2004;2:55. 10.1186/1477-7525-2-55

25. Verstraete J, Ramma L, Jelsma J. Item generation for a proxy health related quality of life measure in very young children. Health Qual Life Outcomes 2020;18:11. 10.1186/s12955-020-1271-1

26. Smith HS, Rubanovich CK, Robinson JO, Levchenko AN, Classen SA, Malek J, et al. Measuring perceived utility of genomic sequencing: Development and validation of the GENEtic Utility (GENE-U) scale for pediatric diagnostic testing. Genet Med 2024;26:101146. 10.1016/j.gim.2024.101146

27. Taylor A, Atkins R, Kumar R, Adams D, Glover V. A new Mother-to-Infant Bonding Scale: links with early maternal mood. Arch Womens Ment Health 2005;8:45–51. 10.1007/s00737-005-0074-z

28. Kerruish NJ, Settle K, Campbell-Stokes P, Taylor BJ. Vulnerable Baby Scale: development and piloting of a questionnaire to measure maternal perceptions of their baby’s vulnerability. J Paediatr Child Health 2005;41:419–23. 10.1111/j.1440-1754.2005.00658.x

29. Brehaut JC, O’Connor AM, Wood TJ, Hack TF, Siminoff L, Gordon E, Feldman-Stewart D. Validation of a decision regret scale. Med Decis Making 2003;23:281–92. 10.1177/0272989×03256005

30. Sanderson SC, Lewis C, Hill M, Peter M, McEntagart M, Gale DP, et al. Decision-making, attitudes and understanding amongst patients and relatives invited to undergo genome sequencing in the 100,000 Genomes Project: a multi-site survey study Genet Med 2022;24:61-74.

31. Braun V, Clarke V. Thematic analysis: A practical guide: SAGE Publications Ltd; 2021.

32. Bradley EH, Curry LA, Devers KJ. Qualitative data analysis for health services research: developing taxonomy, themes, and theory. Health Serv Res 2007;42:1758–72. 10.1111/j.1475-6773.2006.00684.x

33. Genomics England. Your Data in the National Genomic Research Library. https://www.genomicsengland.co.uk/patients-participants/data [

34. World Health O, Stop TBP. Advocacy, communication and social mobilization for TB control: a guide to developing knowledge, attitude and practice surveys. Geneva: World Health Organization; 2008.

35. Weiner BJ, Lewis CC, Stanick C, Powell BJ, Dorsey CN, Clary AS, et al. Psychometric assessment of three newly developed implementation outcome measures. Implement Sci 2017;12:108. 10.1186/s13012-017-0635-3

36. Michie S, Dormandy E, Marteau TM. The multi-dimensional measure of informed choice: a validation study. Patient Educ Couns 2002;48:87–91. 10.1016/s0738-3991(02)00089-7

37. Buchanan J, Hill M, Vass CM, Hammond J, Riedijk S, Klapwijk JE, et al. Factors that impact on women’s decision-making around prenatal genomic tests: An international discrete choice survey. Prenat Diagn 2022;42:934–46. 10.1002/pd.6159

38. Bessey A, Chilcott J, Pandor A, Paisley S. The Cost-Effectiveness of Expanding the UK Newborn Bloodspot Screening Programme to Include Five Additional Inborn Errors of Metabolism. Int J Neonatal Screen 2020;6. 10.3390/ijns6040093

39. Fusco F, Chudleigh J, Holder P, Bonham JR, Southern KW, Simpson A, et al. Delivering Positive Newborn Screening Results: Cost Analysis of Existing Practice versus Innovative, Co-Designed Strategies from the ReSPoND Study. Int J Neonatal Screen 2022;8. 10.3390/ijns8010019

40. Rose AM, Grosse SD, Garcia SP, Bach J, Kleyn M, Simon NE, Prosser LA. The financial and time burden associated with phenylketonuria treatment in the United States. Mol Genet Metab Rep 2019;21:100523. 10.1016/j.ymgmr.2019.100523

41. Gündüz M, Yüksel Güdek Y, Kasapkara Ç S. Out-of-pocket health expenditures in patients living with ınborn errors of metabolism. Orphanet J Rare Dis 2023;18:179. 10.1186/s13023-023-02775-6

42. Desai AD, Zhou C, Stanford S, Haaland W, Varni JW, Mangione-Smith RM. Validity and responsiveness of the pediatric quality of life inventory (PedsQL) 4.0 generic core scales in the pediatric inpatient setting. JAMA Pediatr 2014;168:1114–21. 10.1001/jamapediatrics.2014.1600

43. Varni JW, Limbers CA, Neighbors K, Schulz K, Lieu JE, Heffer RW, et al. The PedsQL™ Infant Scales: feasibility, internal consistency reliability, and validity in healthy and ill infants. Qual Life Res 2011;20:45–55. 10.1007/s11136-010-9730-5

